# Predictive Accuracy of a Hierarchical Logistic Model of Cumulative SARS-CoV-2 Case Growth

**DOI:** 10.1101/2020.06.15.20130989

**Authors:** Levente Kriston

## Abstract

**Background:** Infectious disease predictions models, including virtually all epidemiological models describing the spread of the SARS-CoV-2 pandemic up to June 2020, are rarely evaluated. The aim of the present study was to investigate the predictive accuracy of a prognostic model for forecasting the development of the cumulative number of reported SARS-CoV-2 cases in countries and administrative regions worldwide.

**Methods:** The cumulative number of reported SARS-CoV-2 cases was forecasted in 251 regions with a horizon of two weeks, one month, and two months using a previously described hierarchical logistic model at the end of March 2020. Forecasts were compared to actual observations by using a series of evaluation metrics.

**Results:** On average, predictive accuracy was very high in nearly all regions at the two weeks forecast, high in most regions at the one month forecast, and notable in the majority of the regions at the two months forecast. Higher accuracy was associated with the availability of more data for estimation and with a more pronounced cumulative case growth from the first case to the date of estimation. In some strongly affected regions, cumulative case counts were considerably underestimated.

**Conclusions:** With keeping its limitations in mind, the investigated model can be used for the preparation and distribution of resources during the SARS-CoV-2 pandemic. Future research should primarily address the model’s assumptions and its scope of applicability. In addition, establishing a relationship with known mechanisms and traditional epidemiological models of disease transmission would be desirable.

## BACKGROUND

Mathematical and simulation models of infectious disease dynamics are essential for understanding and forecasting the development of epidemics.^1^ As of June 2020, the ongoing severe acute respiratory syndrome coronavirus 2 (SARS-CoV-2) pandemic has called increased attention to epidemiological modeling both as a method of scientific inquiry and as a tool to inform political decision making.^2–5^

Among epidemiological modeling methods, a distinction between mechanistic and phenomenological approaches is frequently made. While mechanistic approaches model the transmission dynamics based on substantial concepts from biology, virology, infectology, and related disciplines, phenomenological (sometimes termed ‘statistical’) models are looking for a mathematical function that fits observed data well without clear assumptions about the underlying processes.^1,2^ Mechanistic models are usually used to compare possible scenarios and to estimate the relative effects of different interventions rather than to produce precise predictions. On the contrary, phenomenological models are commonly optimized for forecasting. From a broader perspective, mechanistic and phenomenological approaches can be considered as the epidemiological modeling representatives of the long-standing explanation-prediction controversy.^6^ It should be noted that although the distinction between these two model classes is instructive and one side usually predominates, most approaches have both mechanistic and phenomenological components, and some are explicitly balanced (so called ‘semi-mechanistic’ or ‘hybrid’ models).

Although the value of any predictive model is ultimately determined by whether it improved critical decision making,^7,8^ a rigorous scientific appraisal should also include a comparison of what have been predicted to what have actually happened.^1,9,10^ Unfortunately, infectious disease predictions models are rarely evaluated during or after outbreaks.^9,10^ Notable exceptions include systematic evaluation of models about the epidemiology of severe acute respiratory syndrome (SARS),^11,12^ influenza,^13,14^ ebola,^7,9,15,16^ dengue,^10,17^ foot-and-mouth disease,^8^ and trachoma.^18^

The SARS-CoV-2 pandemic has prompted a large amount of epidemiological modeling efforts, including studies with primarily mechanistic (e.g., references^19–25^), primarily phenomenological (e.g., references^26–29^), and hybrid (e.g., reference^30^) approaches. According to the knowledge of the author up to June 2020, none of the existing models was systematically evaluated with regard to the accuracy of their predictions. In order to start closing this gap, the objective of the present study was to evaluate the predictive accuracy of a phenomenologically oriented model that was trained on data up to the end of March 2020 for forecasting the development of the cumulative number of reported SARS-CoV- 2 cases in countries and administrative regions worldwide.^31^

## METHODS

### Data

As described in detail elsewhere,^31^ the model was fitted using information on the cumulative number of confirmed SARS-CoV-2 infections in the COVID-19 data repository of the Johns Hopkins University Center for Systems Science and Engineering.^32,33^ Cumulative case count data from 251 countries and administrative regions were used for training the model, with daily time series from the day of the first reported case to 29 March 2020 in each region. For evaluation, data of confirmed cases were extracted from the same database two weeks, one month, and two months after model development (12 April, 29 April, and 29 May 2020). Sufficient information for creating predictions of the most likely number of cases in all investigated countries and administrative regions for any time horizon was made publicly available at the beginning of April 2020.^31^

### Model

A hierarchical logistic model was fit to observed data.^31^ The logistic part of the model was based on the ecological concept of self-limiting population growth^34^ and used a formulation with five parameters^35^ controlling the expected final case count at the end of the outbreak (parameter *a*), the maximum speed of reaching the expected final case count (parameter *b*), the approximate time point of the transition of the outbreak from an accelerating to a decelerating dynamic (parameter *c*), the case count at the beginning of the outbreak (parameter *d*), and the degree of asymmetry between the accelerating and decelerating phases of the outbreak (parameter *g*). The predicted number of cumulative case counts in region *i* at day *t* from the first reported case was estimated as

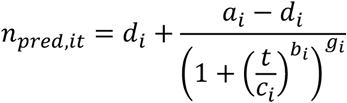

with log-normally distributed errors.

The hierarchical part of the model was inspired by random-effect meta-analysis assuming that the parameters of the logistic equation are similar, but not necessarily identical, across the investigated regions.^36,37^ This was implemented by restricting the parameters of the logistic equation to follow a normal distribution in the population of regions. With respect to interpretation, this means that the model was based on the hypothesis that the pandemic runs a similar course in all countries and regions, even though they are expected to differ to a certain degree regarding the number of cases in their first report, the expected final case count, the time point and speed of the accelerating and decelerating phases of the outbreak, as well as the time point, extent, and effects of control measures.

### Evaluation metrics

For evaluating each individual estimate *i* at time point *t*, four measures were calculated.

The difference between logarithmic predicted and observed counts (“error in logs”, *EIL*) was defined as

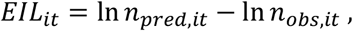

with ln being the natural logarithm, and *n*_*pred*_ and *n*_*obs*_being the predicted and the observed cumulative case counts, respectively.

The absolute error in logs (*AIEL*) was calculated as

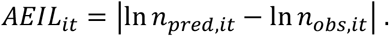

The percentage error (*PE*) was calculated as

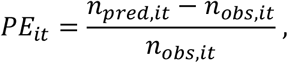

and the absolute percentage error (*APE)* as

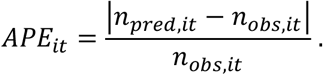

Summary estimates of predictive accuracy across all *k* regions at a given time point *t* are listed in the following.

The root mean squared error in logs (*RMSE*) was defined as

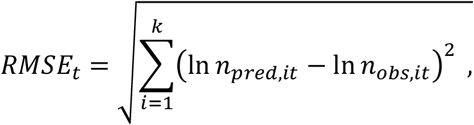

and the mean absolute percentage error (*MAPE*) was calculated as

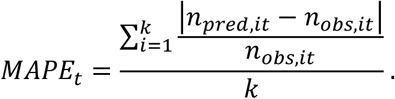

The coefficient of determination *R*^*2*^ _*t*_ was additionally determined from a linear model regressing the logarithmic observed values on the logarithmic predictions with the intercept fixed at zero. Furthermore, the intraclass correlation coefficient *ICC(3,1)*_*t*_was calculated for quantifying the level of absolute agreement between predicted and observed values from a two-way mixed-effects model.^38^ Bootstrapping was used with 1,000 samples to create 95% confidence intervals for summary estimates of predictive accuracy.

### Factors associated with accuracy

In order to identify factors associated with the accuracy of the predictions, the *AEIL* was regressed on the number of available data points, the difference in the logarithm of the first and the last case count at the moment of estimation (as a proxy for progress of the epidemic), and their interaction term. Estimates are reported with 95% parametric confidence intervals.

Furthermore, strongly affected regions (defined by a minimum of 10,000 cases at the forecasted time point) with the most extreme under- and overestimation were identified to gain additional qualitative insights on model performance.

## RESULTS

### Data

In 251 regions, the number of available data points at estimation ranged from 2 to 68 with a median of 25 and a mean of 31.48 days. The cumulative number of reported cases at the point of the first non-zero count ranged from 1 to 444 with a median of 1 and a mean of 4.09 across regions. The cumulative number of reported cases at model estimation (29 March 2020) ranged from 1 to 140,886 with a median of 139 and a mean of 2,869.

### Individual estimates of predictive accuracy

The probability density function of the percentage error (*PE*) at the day of estimation as well at the forecasts after two weeks, one month, and two months, respectively, is displayed in Figure 1. At the day of estimation, the median relative error indicated an average underestimation of the cumulative case count by about one third across regions. The relative error distribution was rather narrow, with only a tenth of predictions showing an underestimation exceeding −62.8 percent and none of the predictions having more than 36.9 percent error. Across forecasts, the median percentage error was always less than twenty percent, although an overestimation by more than two hundred percent was observed in 7.2, 19.1, and 19.5 percent of the cases at the two weeks, one months, and two months forecasts, respectively. The proportion of regions with an underestimation exceeding minus two thirds (−66.6 percent) was 12.4, 19.5, and 28.7 percent at the two weeks, one months, and two months forecasts, respectively.

**Figure 1.**
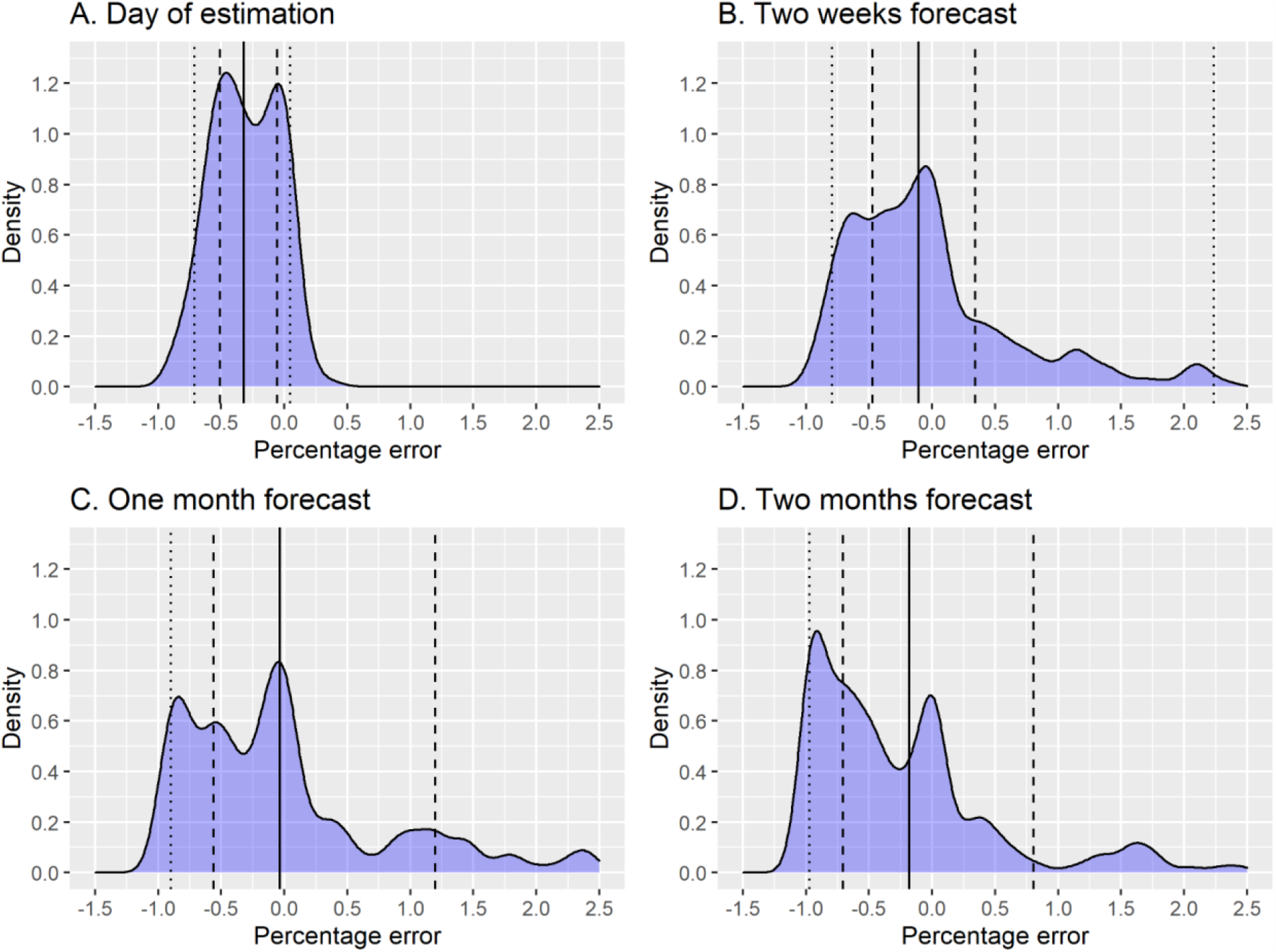
Probability density function of the percentage error at different forecast horizons. The solid line shows the median, the dashed lines show the first and third quartiles, and the dotted lines show the first and ninth deciles. The x-axis is trimmed at 2.5.

The calibration plots suggest an increasing number of regions for which case counts are substantially under- or overestimated with increasing length of the forecast period (Figure 2). Nevertheless, a strong positive association between predicted and observed case counts is apparent even after two months.

**Figure 2.**
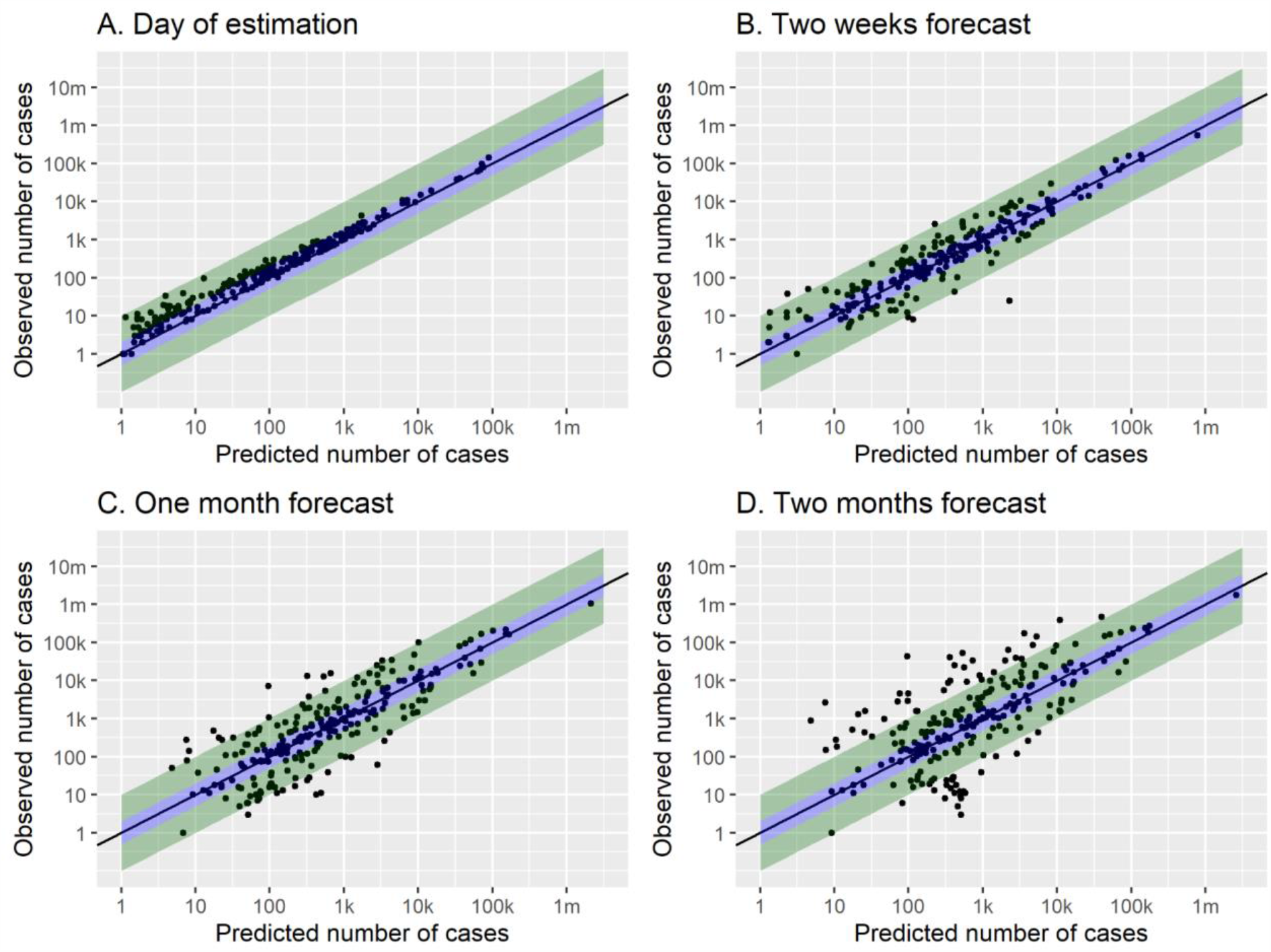
Calibration plots at different forecast horizons. Points refer to regions. The solid black line indicates no prediction error, the blue area indicates a prediction error by a factor of two or less, and the green area indicates a prediction error by a factor of ten or less. Both axes are log-transformed.

### Summary estimates of predictive accuracy

All parameters show an increasing amount of error with increasing length of the forecast period (Table 1). The MAPE shows that, on average, estimates are off by more than one hundred, two hundred, and four hundred percent at the two weeks, one month, and two months forecasts, respectively. The coefficient of determination indicates a very strong relative association between predicted and observed case counts, and the intraclass correlation coefficient suggests that the level of absolute agreement is excellent after two weeks and still high after one month, but sinks to a moderate level after two months.

**Table 1.** Summary estimates of predictive accuracy.

|  | RMSE<br>(95% CI) | MAPE<br>(95% CI) | R<sup>2</sup><br>(95% CI) | ICC<br>(95% CI) |
| --- | --- | --- | --- | --- |
| *Day of estimation* | 0.640<br>(0.577 to 0.707) | 0.323<br>(0.295 to 0.356) | 0.989<br>(0.986 to 0.992) | 0.984<br>(0.979 to 988) |
| *Two weeks forecast* | 0.900<br>(0.803 to 1.05) | 1.085<br>(0.673 to 2.598) | 0.980<br>(0.971 to 0.984) | 0.935<br>(0.905 to 0.950) |
| *One month forecast* | 1.393<br>(1.271 to 1.546) | 2.133<br>(1.600 to 2.953) | 0.958<br>(0.948 to 0.966) | 0.828<br>(0.777 to 0.866) |
| *Two months forecast* | 1.958<br>(1.791 to 2.157) | 4.250<br>(2.907 to 6.735) | 0.931<br>(0.914 to 0.943) | 0.679<br>(0.606 to 0.748) |

RMSE=root mean squared error in logarithmic case counts; MAPE=mean absolute percentage error in case counts; R<sup>2</sup>=coefficient of determination; ICC=intraclass correlation; CI=confidence interval

### Factors associated with accuracy

Visual analysis suggests that a larger number of available data points at estimation (Figure 3) and a more extensive growth of the logarithmic case counts from the first reported case until estimation (Figure 4) are associated with a lower prediction error.

**Figure 3.**
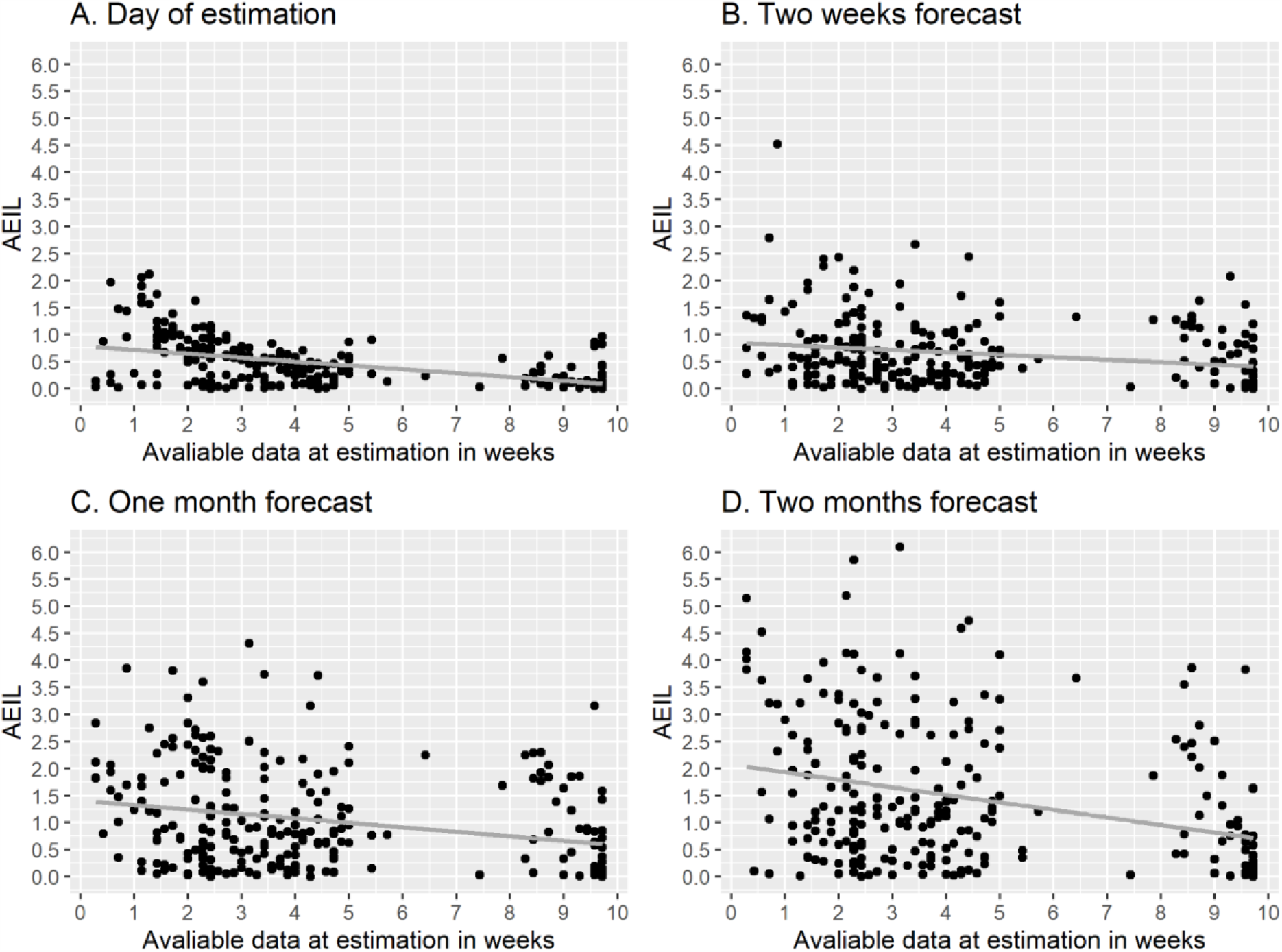
Association of the amount of available data at estimation and predictive accuracy (AEIL) at different forecast horizons. AEIL= absolute difference between logarithmic predicted and observed case counts. Points refer to regions. The grey line corresponds to a linear smoothing curve.

**Figure 4.**
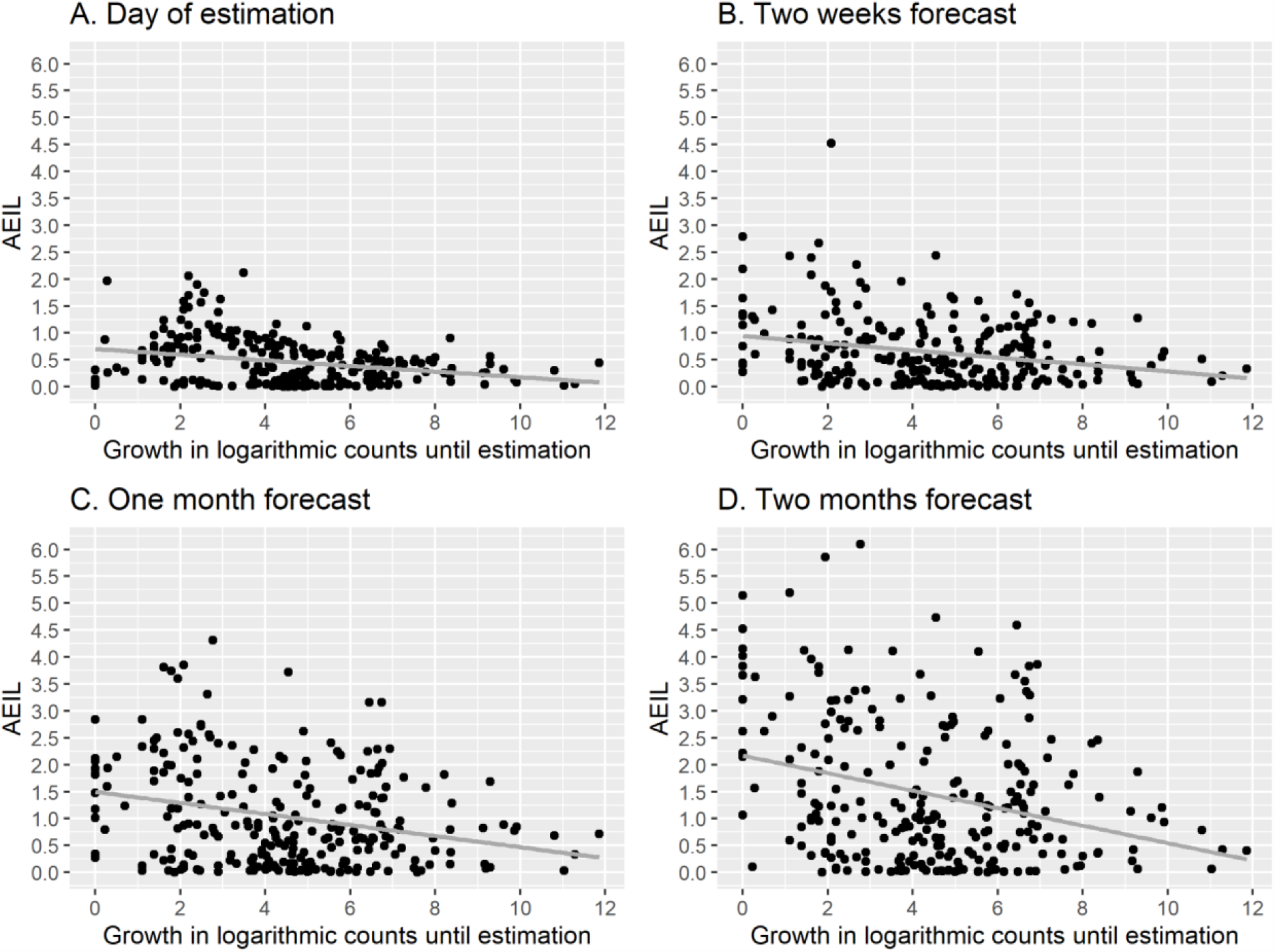
Association of growth in logarithmic case counts until estimation and predictive accuracy (AEIL) at different forecast horizons. AEIL= absolute difference between logarithmic predicted and observed case counts. Points refer to regions. The grey line corresponds to a linear smoothing curve.

This is confirmed by regression analyses indicating statistically significant associations that are becoming stronger with increasing forecast horizon (Table 2). These two factors have also a multiplicative effect, as indicated by the statistically significant interaction term.

**Table 2.** Regression coefficients for factors associated with prediction accuracy (AEIL)

|  | Number of data points in weeks<br>(95% CI) | Growth in logarithmic case counts until estimation<br>(95% CI) | Interaction term<br>(95% CI) |
| --- | --- | --- | --- |
| *Day of estimation* | -0.077***<br>(-0.114 to -0.040) | -0.016<br>(-0.055 to 0.023) | 0.002<br>(-0.005 to 0.009) |
| *Two weeks forecast* | -0.073*<br>(-1.304 to -0.015) | -0.100**<br>(-1.614 to -0.039) | 0.011*<br>(0.000 to 0.022) |
| *One month forecast* | -0.131**<br>(-0.216 to -0.046) | -0.145**<br>(-0.235 to -0.054) | 0.017*<br>(0.001 to 0.034) |
| *Two months forecast* | -0.242***<br>(-0.361 to -0.124) | -0.242***<br>(-0.368 to -0.117) | 0.032**<br>(0.010 to 0.055) |

AEIL= absolute difference between logarithmic predicted and observed case counts; CI=confidence interval; \*p<.050; \*\*p<.010; \*\*\*p<.001

**Table 3.** Most extreme under- or overestimation for regions with a minimum number of 10,000 cases.

|  | Underestimation |  | Overestimation |  |
| --- | --- | --- | --- | --- |
|  | Region | EIL | Region | EIL |
| *Day of estimation* | Belgium | -0.565 | Hubei, China | 0.022 |
|  | United States of America | -0.444 | Germany | 0.020 |
|  | Netherlands | -0.422 | NA | NA |
|  | Switzerland | -0.322 | NA | NA |
|  | Italy | -0.301 | NA | NA |
| *Two weeks forecast* | Belgium | -1.274 | Austria | 0.657 |
|  | Sweden | -1.171 | Quebec, Canada | 0.498 |
|  | Russia | -0.939 | Switzerland | 0.399 |
|  | France | -0.651 | United States of America | 0.336 |
|  | Iran | -0.556 | Germany | 0.096 |
| *One month forecast* | Belarus | -3.719 | Austria | 1.281 |
|  | Qatar | -3.159 | Switzerland | 0.889 |
|  | Singapore | -3.155 | United States of America | 0.714 |
|  | India | -2.301 | Quebec, Canada | 0.638 |
|  | Russia | -2.290 | Portugal | 0.402 |
| *Two months forecast* | Bangladesh | -6.097 | Austria | 1.398 |
|  | Belarus | -4.730 | Switzerland | 1.012 |
|  | Qatar | -4.597 | United States of America | 0.399 |
|  | Kuwait | -4.104 | Israel | 0.358 |
|  | India | -3.864 | Portugal | 0.302 |

EIL= difference between logarithmic predicted and observed case counts; NA=not applicable

Strongly affected regions (a minimum of 10,000 cases) with extreme under- or overestimation of the cumulative case counts are presented in Table 4. Among the listed regions, the extent of underestimation was considerable (an *EIL* below −1.6, roughly corresponding to an underestimation by a factor of five) at the one and two months forecasts, with most regions being located in Asia. Among strongly affected regions, overestimation was rather moderate (an *EIL* below 0.7, roughly corresponding to an overestimation by a factor of two) in most cases. Substantial overestimation (an *EIL* between 0.7 and 1.6) was present in Austria and Switzerland at the one and two months forecasts and in the United States at the one month forecast. No strongly affected region with a considerable overestimation (*EIL* above 1.6) was identified.

## DISCUSSION

In the present study, a hierarchical logistic model was used to predict cumulative counts of confirmed SARS-CoV-2 cases in 251 countries and administrative regions with two weeks, one month, and two months forecasting horizons. Several metrics were used to evaluate predictions visually and statistically. In summary, case counts could be predicted in the majority of the regions with a surprising accuracy. In spite of the facts that at the time of estimation (29 March 2020) only about one month’s data were available on average in each region, and that most regions were at the very beginning of the epidemic, a massive difference between forecast and observation was rather the exception than the rule. Summary metrics of predictive accuracy suggested very strong prognostic validity the model for a horizon of two weeks, substantial accuracy after one month, and still notable, although markedly lower, accuracy after two months. This is in good agreement with studies finding that the horizon for reasonable epidemiological predictions covers a few weeks at most.^9,17^

Although most predictions were fairly accurate, some were still considerably off. They were most likely to be found in regions with a lower amount of available data at the date of estimation and/or with a more limited growth between the date of the first case and the date of estimation. In general, underestimation seems to be somewhat more pronounced than overestimation, particularly in strongly affected regions (i.e., with cumulative case counts above 10,000 at the point of validation). The strongly affected regions for which the model provided too low predictions included several countries in which mitigation strategies might have been less effective than in other regions, as suggested by the only slowly or not at all decelerating cumulative case growth curves at the beginning of June 2020 (e.g., India, Bangladesh, Qatar). On the other hand, the strongly affected regions with a substantial overestimation of cumulative case counts are characterized with an extremely successful mitigation of the epidemic (mainly Austria and Switzerland). Hence, predictive errors are likely to be closely related to one of the central assumptions of the model, i.e., that timing, extent, and effectiveness of control measures is comparable across regions. Obviously, the forecasts based on the presented model are likely to reach their limits in regions that deviate too strongly from the average case. As shifting individual estimates towards the group mean is also a statistical property of hierarchical models,^39^ extreme cases are likely to fall outside the scope of validity of the presented approach.

A notable feature of the model that it provides predictions without any reference to measures taken to control the epidemic. This “ignorance” towards interventions, paired with fairly accurate predictions, may be misinterpreted as evidence of dispensability of the mitigation and containment measures implemented in most countries. However, it is far more likely that the key model assumption suggesting similarity of the course of the epidemic and of the control measures taken across regions holds to a substantial extent. In cases when it does not, model performance is very poor, as discussed above. Bringing these issue together, the hierarchical structure of the model appears to have both benefits and risks: sufficiently accurate predictions for a large number of regions even at a very early stage of the epidemic come with the price of considerably erroneous predictions for atypical regions. Consequently, if used with the aim of generating locally applicable predictions for a particular region, forecasts may be improved by using data from comparable regions with a higher probability than from rather dissimilar regions.^40^

The presented evaluation study has several limitations. First, the case counts were not standardized in any form. Expressing them as cumulative incidence rates (e.g., per 100,000 persons) is likely to have increased homogeneity across regions and enhanced interpretability. As it has been shown in a specific analysis of the development of the SARS-CoV-2 epidemic in German federal states, standardization has rendered using log-transformation of case counts for homogenization superfluous and allowed estimating models with normally distributed errors.^41^ Second, in the present study uncertainty of the predictions remained unconsidered, although measures of uncertainty, such as reliability and sharpness, can be just as important for forecasting as bias.^9^ Third, predictions only at selected time points were analyzed, and it cannot be excluded that choosing other time points would have led to different results. Nevertheless, the general pattern of findings is unlikely to have changed substantially. The forecasting model itself has some weaknesses as well.^31,41^ Most importantly, it models the reported rather than the true number of cases and therefore can be subject to different forms of testing and reporting bias. Considerable improvement regarding this point can realistically be expected first when regional findings form well-conducted epidemiological studies become available. Another major limitation of the model is that it works only as long as the conditions of the epidemic remain largely unchanged in each region, i.e., within a single epidemic wave with fairly constant testing and reporting practices and without serious disruptions. This issue could perhaps be addressed by using dynamic (time-dependent) rather than fixed (time-invariant) model parameters.^42^ Finally, the primarily phenomenological nature of the model calls for integration with mechanistic components, in order to create a hybrid approach that is capable of producing widely generalizable conclusions.^43^

As stated by one of the most prominent epidemiologist of the SARS-CoV-2 pandemic, Neil Ferguson, models are “not crystal balls”.^3^ However, without rigorous scientific evaluation, they run the risk of becoming one, characterized not by correct predictions but by obscurity. Some state that epidemiological forecasting is “more challenging than weather forecasting”,^44^ and complexity of modelling and reliance on assumptions make it difficult to assess the trustworthiness of models based solely on their inherent structure. Just like we trust weather forecasts that prove to be accurate by experience, empirical comparison of modeling predictions with actual observations should become an essential step of epidemiological model evaluation.

## Data Availability

The analyzed data are publicly available.

## Acknowledgments

The study was not externally funded.

## Conflict of interest

The author reports no conflict of interest.

## Data availability

The analyzed data are publicly available.

## Notes

### Competing Interest Statement

The authors have declared no competing interest.

